# Patients with Type 2 Diabetes and a *GCK* variant are still at risk for T2D-related secondary complications

**DOI:** 10.1101/2023.12.19.23300223

**Authors:** Kelly M. Schiabor Barrett, Natalie Telis, Lisa M. McEwen, Evanette K. Burrows, Daniel P. Judge, Pamala A. Pawloski, Joseph J. Grzymski, Nicole L. Washington, Alexandre Bolze, Elizabeth T. Cirulli

## Abstract

Natural HbA1c levels in *GCK*-MODY patients often sit above the diagnostic threshold for type 2 diabetes (T2D). Standard treatments to lower HbA1c levels are ineffective in these individuals, yet in case studies to date, *GCK*-MODY patients often evade secondary T2D complications. Given these deviations from a more typical T2D disease course, genetic screening of *GCK* may be clinically useful, but population studies are needed to more precisely quantify T2D-related outcomes in *GCK* variant carriers. Using a state-of-the-art variant interpretation strategy based on glucose elevations, we genotyped all individuals in two real-world cohorts (n~535,000) for *GCK* risk variants and examined rates of T2D and T2D-complications from seven disease categories.

We identified 439 individuals with *GCK* variants predicted to increase glucose (~1/1200). Aligning with their glucose elevations, *GCK*-MODY variant carriers were 12x as likely, and all other *GCK* risk carriers 4x as likely, to receive a T2D diagnosis, compared to non-*GCK* carriers. Surprisingly, *GCK* risk carriers with T2D develop a range of T2D-related complications at rates comparable to non-*GCK* T2D patients. Although the penetrance for secondary complications is lower than that for glucose elevations, *GCK* risk carriers remain at elevated risk of T2D and secondary complications.

---

Type 2 Diabetes (T2D) is a common, complex disease affecting 1 in 10 Americans. It is often a diagnosis of exclusion centered on elevations in a single biomarker: blood glucose or its close derivative, glycated hemoglobin (HbA1c), and it is typically diagnosed in adulthood, after age 45^1^. Independently, the gene *GCK* is also closely tied to blood glucose levels. *GCK* encodes for a hexokinase expressed largely in pancreatic beta cells and liver hepatocytes. *GCK* gates glucose and insulin regulation through glucose sensing and processing, ultimately playing a key role in setting and maintaining glucose homeostasis for the body. Both rare loss of function and a subset of damaging missense variants in *GCK* have been shown to dampen or reduce the enzymatic functions of the protein, resulting in an elevated endogenous glucose set point which, on average, sits just above the clinical diagnosis threshold for T2D ^2–5^.

Clinically-recognized glucose elevations amongst individuals harboring damaging *GCK* variants have near complete penetrance, although most individuals harboring these variants are unaware of their genotype and its association with plasma glucose levels ^6–9^. If elevated glucose is first measured in childhood or early adulthood, a “maturity onset diabetes of the young” (MODY) diagnosis and a genetic etiology, such as *GCK*, may be considered ^10^. If first measured in adulthood, a clinical T2D diagnosis is more likely to be assigned, as monogenic etiologies are not typically suspected or tested for in this age range ^11,12^. Strong associations have been observed between individuals harboring *GCK* variants and clinical T2D diagnoses in both T2D-specific cohorts, where up to 1% of enrolled patients harbor damaging *GCK* variants, as well as in gene-disease association studies of all-comer, clinico-genomic cohorts ^8,12–17^.

Genetic screening of *GCK* has the potential to be clinically useful. Penetrance estimates for *GCK* and T2D are in line with some of the most well established CDC Tier-1 gene-disease associations, such as *BRCA1*/*BRCA2* with Breast and Ovarian Cancers and *LDLR*/*APOB* with Familial Hypercholesterolemia^13^. Early work focused on outcomes suggests that those harboring *GCK* variants and standalone mild, stable hyperglycemia are not at increased risk for the typical array of long-range complications associated with T2D, especially microvascular complications ^14,18–22^. In addition, treating patients with damaging *GCK* variants with first-line pharmacological interventions such as insulin or oral hypoglycaemic agents (metformin and glibenclamide) has been shown to have little to no effect on HbA1c levels, and it may be difficult or impossible to reduce glucose levels below standard risk thresholds ^18,23^. Taken together, the excess of T2D diagnoses in individuals harboring damaging *GCK* variants may represent some degree of misdiagnosis in adulthood of what are instead asymptomatic, mild and stable hyperglycemias in these individuals ^11,18,24^. Untangling benign glucose elevations from accurately assigned T2D diagnoses in those with *GCK* variants represents a great opportunity for precision medicine ^12^. The deviations in both disease progression and treatment response suggest that stand alone natural glucose elevations should be considered alongside a clinical T2D diagnosis. To date, outcomes-related research in those harboring *GCK* variants to date has been limited to comparisons between MODY subtypes, has focused only on those with isolated mild, stable hyperglycemia, has been in the form of case studies and small cohorts, and has largely been limited to clinically described variants ^9,17,19^.

To formulate genetic screening-based diagnosis and treatment recommendations, a comprehensive genotyping strategy to identify and interpret not only loss of function, but also all damaging missense variation across *GCK*, paired with an understanding of the long term T2D-related complication risk of these carriers compared to population controls is needed. The aim of this study is to: 1) develop and validate a population screening variant annotation strategy, with a focus on missense variants, leveraging the highly penetrant elevations in glucose and Hba1c level as a readout, and 2) characterize differences in T2D diagnosis rates, disease progression in terms of secondary outcomes, and the alignment between these diagnoses and long-term outcomes in those carrying qualifying *GCK* variants in multiple large, unselected, adult population cohorts.

## Results

### A population screening variant annotation strategy for *GCK* identifies individuals with high glucose compared to population averages

#### Annotation strategy and carrier prevalence by cohort

We first set out to build a population screening variant annotation strategy to identify individuals with relevant rare (MAF<0.1%) variants in *GCK* that may elevate glucose levels. Aside from variants with a molecular consequence consistent with LoF (LoF), to score all other coding variants for this phenotype we combined the output from two orthogonal datasets that predict glucose elevations for variation across *GCK*. The first dataset is a functional evidence map, which evaluates the functional consequence of all missense and nonsense single nucleotide variants via a yeast complementation assay. In this assay, resulting *GCK* enzymatic activity levels can be quantified by the growth rate of yeast cells carrying each variant on a glucose-based medium ^25^. The second dataset is a statistical evidence map, which evaluates association patterns with glucose levels across the gene using clinico-genomic data, processed by creating statistical power-based windows of rare variants of relevant molecular consequences across a population ^26^. We built the *GCK* statistical evidence map with genetic data from 333,190 randomly selected UKB participants, using random blood glucose (i.e., not necessarily measured after fasting) collected as part of their baseline assessment visit as our phenotype (**Figure 1A**). Leveraging these two resources, we formed a variant annotation strategy using the *GCK* genotypes and, as output, the random blood glucose from the remaining UKB participants (n=112,015, “replication subset”). We found that coding variants that increase glucose levels above the median values of those without *GCK* variants (89 mg/dL) could be reliably identified if: (i) the statistical evidence map predicted the variant to increase glucose levels and (ii) the functional evidence map did not predict the variant to decrease glucose levels. Those with *GCK* variants that did not meet this criteria, on average, displayed glucose levels in line with those without *GCK* variants (**Figure 1B**) ^26,27^.

**Figure 1.**
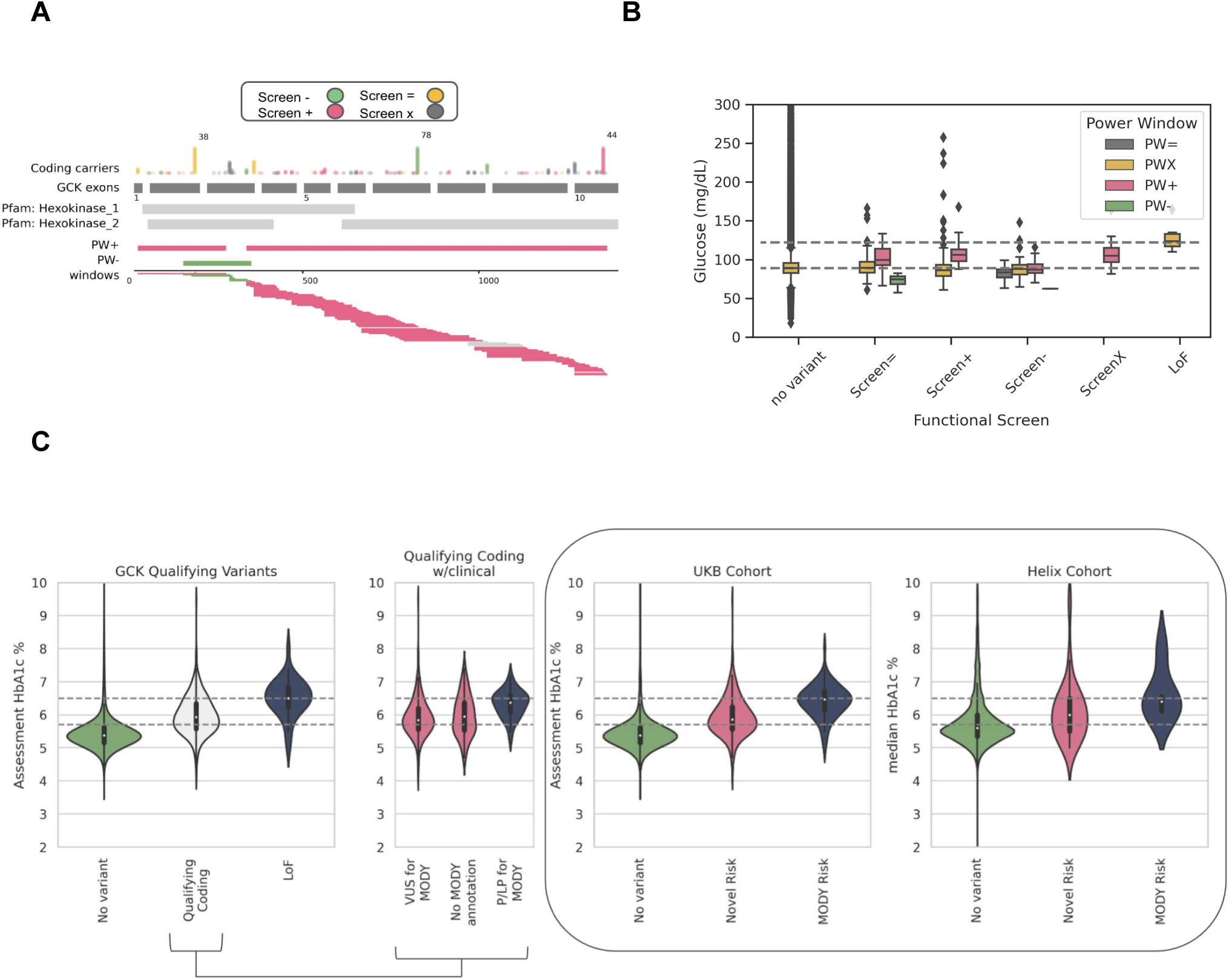
Identifying individuals with qualifying *GCK* variants in the population. **A**) *GCK* statistical evidence map for random blood glucose, created using Power Window (PW), with PW+ regions marked in pink and PW-in green. Variants are plotted by position and colored according to their functional score annotation (Screen+ in pink (functional score <0.66), Screen-in green (functional score >1.18), Screen= in gray (functional score between 0.66 and 1.18), and ScreenX in yellow (no results available from functional screen)) See Methods for details on Power Window technique. **B**) Comparison of random blood glucose levels in individuals with and without *GCK* variants. Individuals are categorized according to whether their *GCK* variant was predicted by the functional evidence map to cause high glucose levels as in A. Individuals with LoF variants or no variants are shown separately. The dashed lines indicate the median values for those without *GCK* variants (89 mg/dL) or those with LoF variants (122 mg/dL). The colors indicate Power Window results: pink (PW+) is those with variants predicted to cause high glucose levels, green (PW-) for low glucose, gray (PW=) for no clear signal, and yellow (PWX) for variants that did not qualify for PW analysis, meaning variants that were MAF>0.1% or were not predicted by Sift or Polyphen to be damaging. The dataset shown is restricted to the 117k UKB individuals used to test PW. For brevity, the plot y axis was cutoff at 300 mg/dL, which excludes 163 individuals from the ‘no variant’ category and 1 from the ‘Screen=’ category. **C**) Assessment and median HbA1c levels by *GCK* variant group, for the subset of individuals with these readings in UKB (n=432,826) and Helix (n=21,744) cohorts, respectively. The first two panels focus on participants from the UKB cohort. The first panel shows qualifying coding and LoF carriers separately, the second panel separates qualifying coding carriers by available MODY annotations; with the HbA1c levels of those with P/LP variants significantly higher than individuals with either a VUS or a variant without annotation, and in line with levels in those with LoF variants. The final panel displays HbA1c levels of genotypic groups used for analyses in this work. Individuals with P/LP qualifying coding and LoF variants are combined for the MODY risk group and individuals harboring qualifying coding variants with either a VUS annotation or without annotation are combined for the Novel risk group. The median HbA1c levels for these groups in UKB were 5.4% for non-carriers, 5.9% for Novel risk group, and 6.5% for MODY risk group; in Helix cohorts they were 5.6% for non-carriers, 6.0% for Novel risk group, and 6.4% for MODY risk group. For brevity, the following individuals were excluded from their respective plots: 995 UKB and 207 Helix participants with HbA1c values >=10%. Gray lines for prediabetic (5.7%) and diabetic (6.5%) clinical HbA1c thresholds are shown for reference. Ses **Figure S1** for Glucose measures by genotype.

Next, we applied our annotation strategy across all individuals in the UKB and Helix cohorts, identifying 333 individuals in UKB and 62 in Helix cohorts who were heterozygous and no homozygotes for qualifying missense or inframe indel coding variants in *GCK*. This represents ~0.07% (1/1400) of UKB and ~0.1% (1/960) of Helix cohorts, respectively or ~50% (333/692 in UKB and 62/128 in Helix) of all individuals with rare, damaging *GCK* coding variants present in each cohort. Additionally, there were 41 individuals in UKB and 3 in Helix cohorts with *GCK* LoF variants. In total, we identified 374 individuals in UKB and 65 in Helix cohorts with qualifying LoF or coding variants in *GCK* (0.082% or 1/1200 in UKB and 0.10% or 1/960 in Helix). This is higher than the previously reported population prevalence for UKB (0.042%) and another American cohort (0.024%), where coding variant interpretation was limited to only previously reported clinical variants and LoF ^8^. Indeed, while the vast majority of clinically-established pathogenic or likely pathogenic (P/LP) variants documented in Clinvar were called as qualifying variants using our annotation strategy (18/20 in UKB and 6/8 in Helix), carriers of this group of variants combined with all individuals harboring LoFs, only represent ~20% of all qualifying variant carriers found using our annotation strategy (83/374 in UKB and 10/68 in Helix). Carrier counts, available functional and statistical data, and basic variant effect information for all P/LP variants in both cohorts, including the 3 variants (one of which was observed in both cohorts) which were not called as qualifying variants using our annotation strategy, are available in **Table S2**. The current age, average age of T2D diagnosis, and a breakdown by sex and genetic similarity for individuals with or without a qualifying variant are presented in **Table 1**.

**Table 1.** Study population demographics.

|  | UKB |  |  | Helix |  |  |  |  |  |
| --- | --- | --- | --- | --- | --- | --- | --- | --- | --- |
|  | No qualifying variant | Coding | LoF | No qualifying variant | Coding | LoF | Cohort 1 | Cohort 2 | Cohort 3 |
| N (%) | 453,564 (99.9%) | 333 (0.07%) | 41 (0.01%) | 62,548 (99.9%) | 62 (0.099%) | 3 (0.004%) | 37,989 | 15,104 | 9,313 |
| P/LP for GCK MODY (%) | 2/453,564 | 42/333 (13%) | 35/41(86%) | 3/62,548 | 6/62(9.7%) | 1/3 (33%) | – | – | – |
| Mean current age (SD) | 70 (8) | 70 (8) | 70 (8) | 54 (17) | 48 (17) | 59 (4) | 54 (17) | 54 (16) | 52 (16) |
| N female (%) | 246,604 (54%) | 161 (54%) | 22 (54%) | 43,270 (69%) | 57 (75%) | 2 (66%) | 25,643 (68%) | 10,915 (72%) | 6,771 (73%) |
| N genetic similarity (%) |  |  |  |  |  |  |  |  |  |
| African | 7,719 (1%) | 0 | 0 | 2,073 (3%) | 2 (3%) | 0 | 746 (2%) | 380 (3%) | 949 (10%) |
| Americas | 0 | 0 | 0 | 6,080 (10%) | 7 (9%) | 0 | 5,331 (14%) | 502 (3%) | 254 (3%) |
| East Asian | 2,989 (1%) | 2 (1%) | 0 | 1,586 (3%) | 6 (8%) | 0 | 1,203 (3%) | 310 (2%) | 79 (1%) |
| South Asian | 7,085 (2%) | 1 (<1%) | 1 (2%) | 393 (1%) | 0 | 0 | 221 (1%) | 123 (1%) | 49 (1%) |
| European | 428,070 (94%) | 286 (98%) | 38 (93%) | 50,320 (81%) | 56 (74%) | 3 (100%) | 29,131 (77%) | 13,474 (89%) | 7,774 (83%) |
| Other | 7,741 (2%) | 4 (1%) | 2 (5%) | 1,875 (3%) | 5 (7%) | 0 | 1,357 (4%) | 315 (2%) | 208 (2%) |
| N with T2D diagnosis (%) | 40,303 (9%) | 82 (28%) | 29 (71%) | 7,210 (12%) | 25 (33%) | 2 (67%) | 4,839 (13%) | 1,437 (10%) | 961 (10%) |
| N with T2D diagnosis prior to baseline (%) | 11,799 (3%) | 33 (11%) | 12 (29%) | NA | NA | NA | NA | NA | NA |
| Mean age of T2D diagnosis (SD) | 62 (9) | 59 (11) | 60 (9) | 58 (14) | 50 (14) | 55 (2) | 59 (14) | 57 (12) | 56 (13) |
| N with glucose measurements (%) | 397,574 (88%) | 252 (86%) | 32 (78%) | 42,056 (67%) | 49 (64%) | 2 (67%) | 32,467 (85%) | 10,069 (67%) | 6,912 (74%) |
| N with HbA1c measurements (%) | 432,498 (95%) | 289 (99%) | 39 (95%) | 20,958 (37%) | 31 (41%) | 1 (33%) | 17,105 (45%) | 1317 (9%) | 3,321 (36%) |

#### Glucose and HbA1c levels in *GCK* qualifying variant carriers are elevated, often into clinically relevant ranges

Across the UKB cohort, we find blood glucose and HbA1c levels are higher for those harboring qualifying *GCK* variants compared to individuals without *GCK* variants, with values highest in those with LoF variants (e.g., the mean random glucose values in those with LoF variants in the UKB was 121 mg/dL, compared to 106 mg/dL for those with qualifying coding variants and 89 mg/dL for those without variants; see **Figure 1C**, **Figure S1 for glucose**). Further, we used standard clinical cutoffs for random glucose and/or HbA1c to classify individuals as either pre-diabetic (HbA1c>=5.7%) or diabetic (HbA1c>=6.5% or random glucose above 200 mg/dL). For UKB, 66% of those with qualifying coding variants and 92% of those with LoF variants had values in the prediabetes range compared to 21% of those without qualifying *GCK* variants. Additionally, 20% of those with coding variants and 51% of those with LoF variants had values in the diabetes range, compared to 4% of those without *GCK* variants (**Figure 1C, S1 for glucose**). We further examined a subset of the UKB cohort who did not have a T2D diagnosis and had not reported taking common T2D medications, insulin or metformin, at the baseline assessment (subcohort counts: LoF=19, qualifying coding=239, no variant= 371,040, **Figure S2**). Elevated glucose and HbA1c trends for those harboring qualifying *GCK* variants held within this undiagnosed subcohort, with 62% of those with coding variants and 90% of those with LoF variants in the pre-diabetic range (HbA1c>=5.7%), compared to 16% of those without qualifying *GCK* variants.

Next, we subsetted individuals harboring qualifying coding variants by presence or absence of a clinical pathogenicity annotation (42 with pathogenic or likely pathogenic (P/LP) annotations, 216 with variant of uncertain significance (VUS), and 75 with no clinical entry). As a subcohort, individuals harboring P/LP qualifying coding variants had mean random glucose (117 mg/dL) and HbA1c (6.4%) levels in line with average levels in LoF carriers and significantly higher compared to the averages for those with either VUS or no clinical annotation (p =0.03 vs. VUS and p=0.005 vs. no clinical annotation by linear regression and **Figure 1C**). Given the close alignment in these relevant lab values and because P/LP missense and LoF variants are not specifically differentiated in clinical interpretation guidelines for MODY^28^, we grouped together individuals harboring either P/LP qualifying coding variants or LoF variants (MODY group, n=83) and evaluated this group against those with VUS qualifying coding variants or qualifying coding variants with no clinical entry (Novel group, n=291), and those without a qualifying variant (No *GCK* Variant, n=453,564) for the remainder of the study (**Figure 1C**).

For a large proportion of the Helix cohorts, random glucose or HbA1c measurements were available from the medical records, taken throughout the course of medical care and often at multiple timepoints. We expected that, unlike the UKB cohort where the measurements were taken for every participant at a single point in time regardless of health history, we would observe higher measurement values due to an enrichment for individuals at risk for diabetes with any measurement. Indeed, taking the max value observed for an individual during their medical history, and the same cutoffs as above, 74% and 31% of those with qualifying coding variants and glucose or HbA1c measurements were pre-diabetic or diabetic, respectively, while 55% and 13% of those without variants were in the prediabetes or diabetes range, respectively. There were three individuals with LoF variants, and of the two with HbA1c or blood measurements, one was in the diabetes range. The mean random glucose values in those with qualifying coding variants in the Helix cohorts was 103 mg/dL, compared to 95 mg/dL for those without variants. Aligning with trends in the UKB cohort, mean random glucose values for MODY(n=10), Novel (n=55), and No *GCK* Variant (n=62,341) groups were: 117, 105, and 95 mg/L, respectively (**Figure 1C, Figure S1 for glucose**).

### Individuals harboring either MODY or Novel qualifying *GCK* variants are at elevated risk for receiving T2D diagnosis at any given time

Overall rates of T2D diagnosis differed by cohort (9% in UKB vs 12% in Helix Cohorts, p=1.3e-293 by logistic regression after controlling for age, sex, BMI, **Table 1**). We next compared diagnostic rates of T2D more specifically for those harboring either MODY or Novel qualifying *GCK* variants compared to those without a qualifying *GCK* variant, using a time to event analysis with all available EHR data points. Despite varying population-level rates of T2D, we find similar T2D diagnosis enrichment between variant groups in both cohorts. Compared to those without *GCK* variants, individuals harboring Novel qualifying variants are approximately 3-4 times more likely to be diagnosed with T2D at any given time (HR=2.8, p=2.74e-16 for UKB and HR=3.7, p=9.32e-8 for Helix) and those harboring MODY qualifying variants are roughly 12 times more likely to be diagnosed with T2D at any given time (HR=12.4, p=1.32e-80 for UKB; Helix HR not possible due to low n but 11x enrichment at age 60, p=2.21e-51 by log rank test for difference between the survival curves).

While MODY is typically diagnosed in childhood or early adulthood, with glucose elevations present at birth, the majority of T2D diagnoses in individuals harboring MODY qualifying variants (49/57 in UKB and 4/7 in Helix) and those with Novel qualifying variants (59/63 in UKB and 12/17 in Helix) were first made after age 45. After removing all T2D diagnoses made prior to 45 years of age, we observed a small, significant earlier age of diagnosis for those with either MODY or Novel qualifying *GCK* variants compared to those without a qualifying *GCK* coding variant in UKB and for those with MODY qualifying variants in Helix cohorts (UKB: MODY 3 years, p=0.001 and Novel 2 years, p=0.02; Helix: MODY 6 years, p=0.07 by linear regression controlling for age, sex, BMI; **Figure 2B**). Finally, in terms of clinical risk stratification, by age 70, in UKB, 79% of individuals carrying MODY variants and 27% of individuals carrying Novel qualifying variants have a T2D diagnosis. In Helix, 100% of MODY carriers and 55% of Novel qualifying variant carriers have a T2D diagnosis. Rates for all carrier groups exceed their relevant population averages (12% for UKB and 20% for Helix by age 70) (**Figure 2A**).

**Figure 2.**
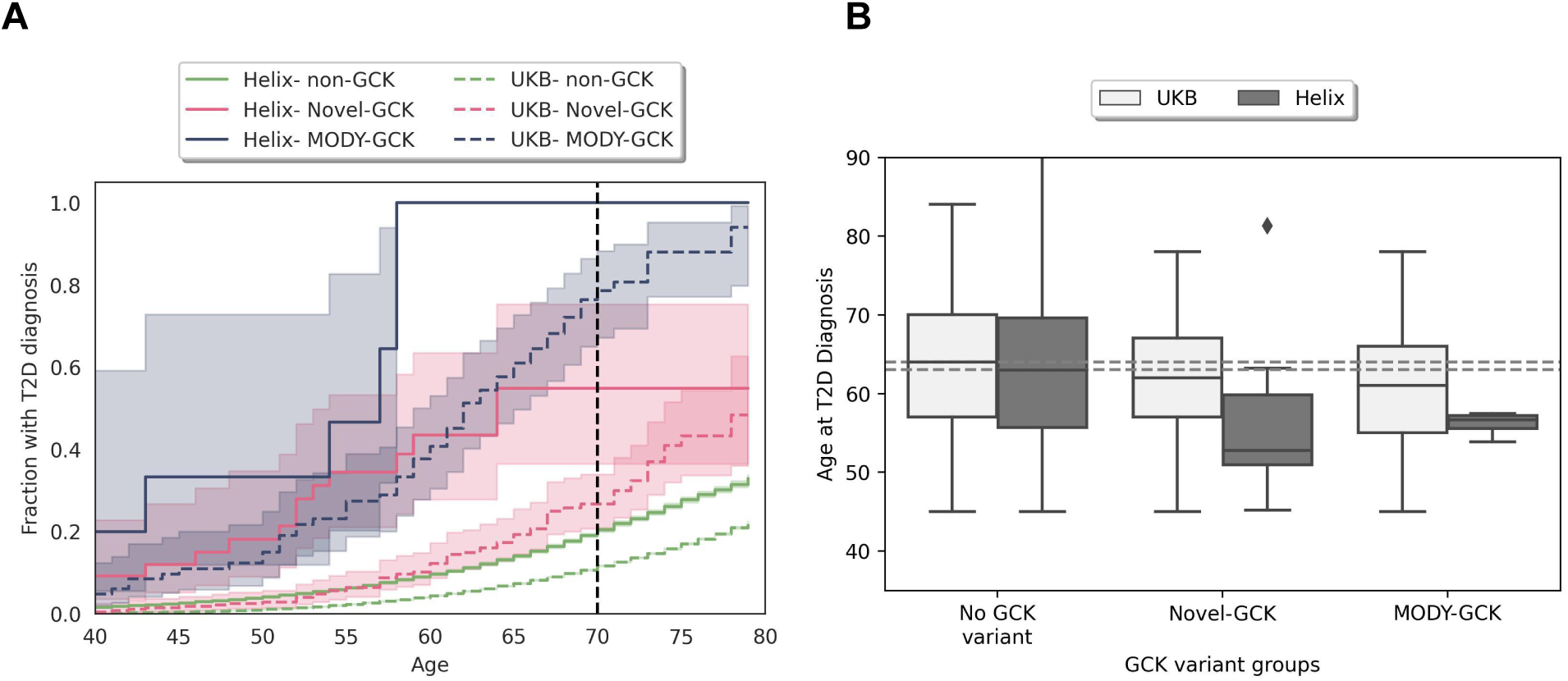
T2D diagnosis prevalence differs by genotype in two cohorts. **A**) Time to event plot for T2D diagnosis by genotype (No variant in green, Novel Risk in pink, and MODY Risk in blue) in UKB Full Cohort (dotted lines) and Helix cohorts (solid lines). For those with Novel Risk variants vs. without, HR=3.8, p=5.8e-34 for UKB and HR=4.3, p=2.7e-13 for Helix. For those with MODY Risk variants vs. without HR=12.4, p=1.32e-80 for UKB 11x enrichment by age 60 Helix.Hazard ratios for subset of cohort with both BMI and sex available (n=452,319 for UKB and n=45,060 in Helix): Those with Novel Risk variants vs. without, HR=3.2, p=5.6e-20 for UKB and HR=3.3, p=3.5e-13 for Helix and those with MODY Risk variants vs. without, HR=14.2, p=1.6e-89 for UKB. **B**) Box plot of age at T2D diagnosis made after age 45 by genotype for UKB (light gray) and Helix (dark gray) cohorts. Diagnosis after age 45 makes up 90% of the UKB diagnoses and 67% of the Helix ones. Median diagnosis age is marked as dotted line age=64 for UKB and age=63 for Helix. Focusing on diagnoses made after age 45, T2D diagnosis happens earlier in those with Novel and MODY qualifying variants (UKB: MODY 3 years, p=0.001 and Novel 2 years, p=0.02; Helix: MODY 6 years, p=0.07 by linear regression).

### DCSI as a tool to quantify T2D outcomes from EHR records

While glucose and HbA1c are linked to T2D diagnosis, these measures are an incomplete representation of clinically significant outcomes themselves. Research suggests not all individuals harboring pathogenic *GCK* variants are truly diabetic in the sense of their likelihood to develop microvascular and macrovascular outcomes ^12,18,19^. In order to assess whether individuals with qualifying MODY or Novel *GCK* qualifying variants diagnosed with T2D had different outcomes compared to other T2D patients, we used the adapted Diabetes Complication Severity Index (DCSI), a 14-point scale, derived from the ICD codes obtained from patient medical records, organized into seven disease categories that are considered secondary complications of T2D (see methods for details, **Table S1** for ICD codes and severity scores for each category) ^29,30^.

We first assessed DCSI scores in all participants of the UKB and Helix cohorts. We find that DCSI captures T2D-related outcomes from available medical records in both cohorts. Average DCSI scores are higher in Helix cohorts compared to UKB, aligning with the differing population rates of T2D (mean DCSI Helix= 0.98; mean DCSI UKB =0.69, for T2D rates see: **Figure 2A**, **Table 1**). Despite differences in mean DCSI in each cohort, DCSI scores increase with age across all participants in each cohort, with the highest scores being held almost exclusively by diabetic patients, as expected for a cumulative score of progressive disease (**Figure 3A,B, S3A,B**). Visualized as time to event curves, accumulation of substantive DCSI scores (DCSI > 3) follow with recorded age of onset of T2D diagnosis, as well as with the number of years since diagnosis for those with T2D (**Figure 3C, S4C**). For our analyses, we assess both cumulative DCSI scores for each individual as well as a dichotomous cutoff of a DCSI of three or more for time to event analyses, which can only be obtained once someone is harboring a diagnosis from at least 2 complication categories (see methods for details).

**Figure 3.**
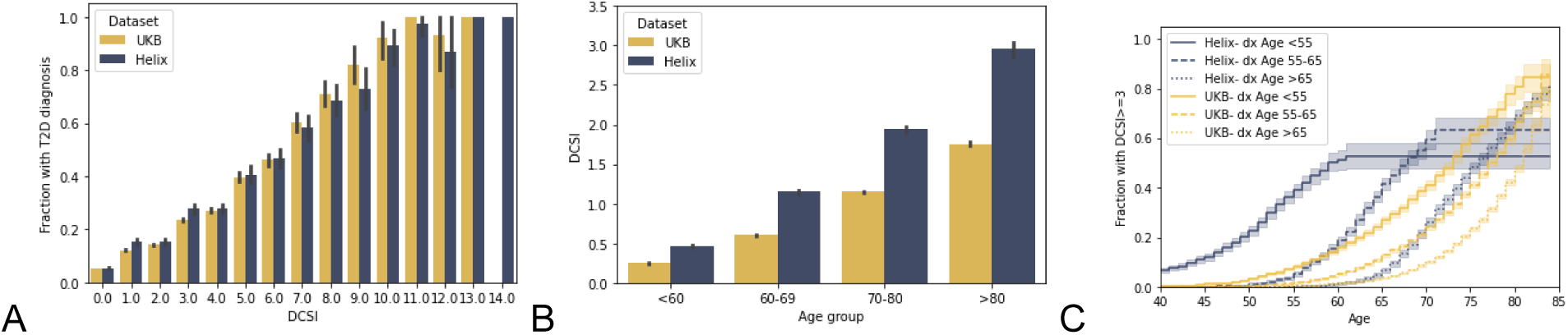
Diabetic Complications Severity Index (DCSI) in UKB and Helix cohorts. T2D is associated with higher DCSI scores. A) Fraction of individuals with a T2D diagnosis for each possible DCSI value (0 to 14). B) DCSI increases with age (see also Figure S4A,B, separated by T2D). Mean DCSI value by age group, calculated across all participants (n=453,938 in UKB; n=62,406 in Helix cohorts). C) Time to event curves for substantial DCSI outcomes (DCSI >3) in T2D cases, bucketed by age at diagnosis (< 55, 55-65, >65). Substantial DCSI outcomes begin to accumulate earlier for those with earlier diagnosis.

### T2D cases with MODY and Novel qualifying GCK variants develop T2D complications at rates similar to the T2D population at large

To determine if MODY and Novel *GCK* qualifying variant carriers with T2D diagnoses are also at some level of elevated risk for T2D-related complications, we performed a time to event analysis, controlling for sex and BMI, and using age at DCSI of three or more as an endpoint. For both variant groups, those with a T2D diagnosis are significantly more likely to develop more than one category of T2D complications as measured by DCSI than were population controls without a T2D diagnosis (MODY UKB (n=57) HR=1.9, p=0.07; MODY Helix (n=7) HR=3.9, p=0.17 and Novel UKB (n=63) HR=2.6, p=0.0006; Novel Helix (n=17) HR=3.2, p=0.01) and accumulate complications at a rate similar to that of other T2D cases (non-*GCK* UKB HR=2.45; Helix HR=3.3, **Figure 4A-B**). Conversely, those with Novel qualifying *GCK* variants who did not receive a T2D diagnosis had a rate of complications in line with that of population controls without T2D (UKB (n=228) HR= 0.81, p=0.54; Helix (n=38) HR 0.97, p=0.97, **Figure S4A-B**). Due to sample size limitations, we did not assess the rate of DCSI complications in those with MODY variants and no T2D diagnosis (UKB n=26, Helix n=3).

**Figure 4.**
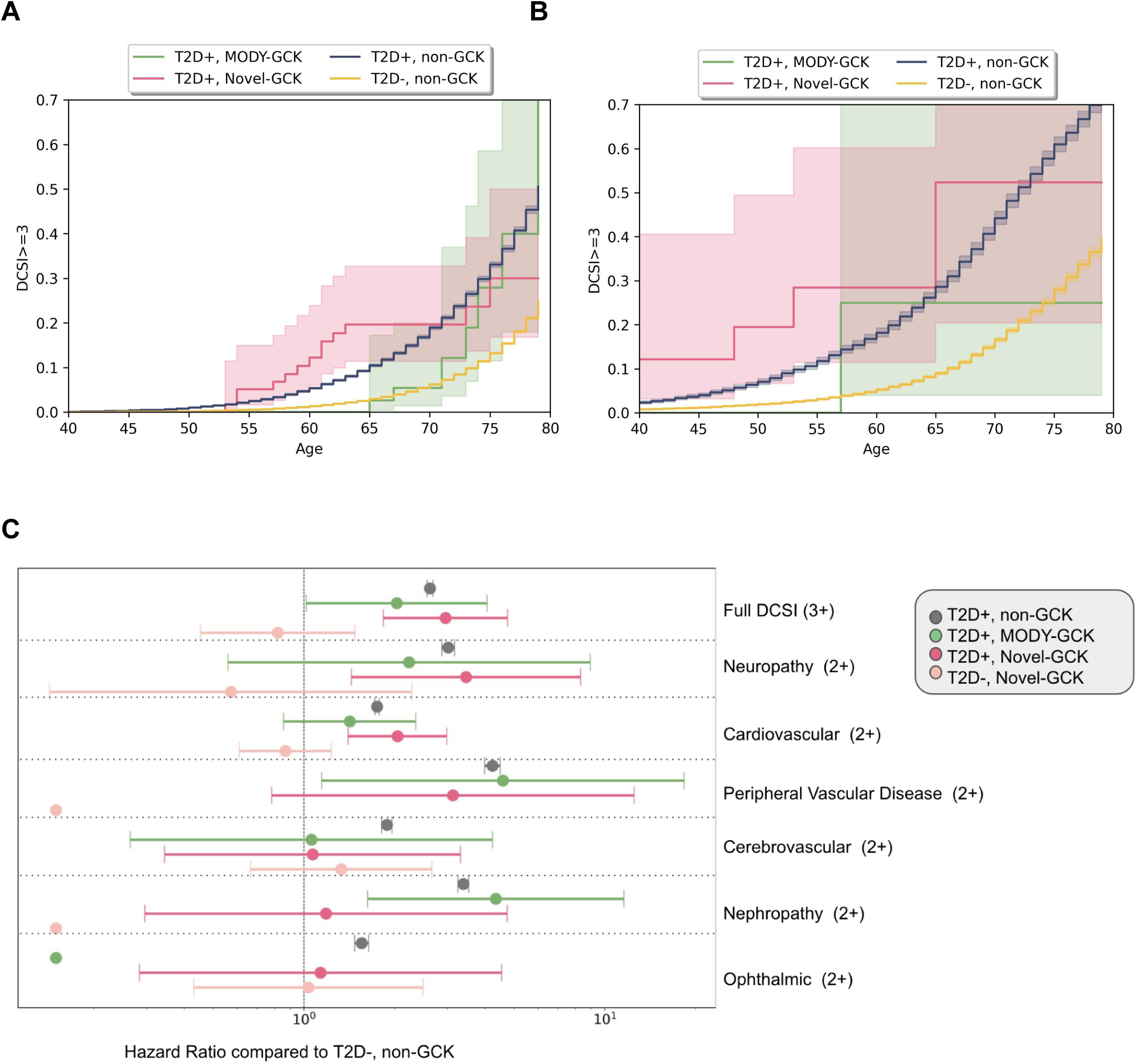
T2D diagnoses are associated with the accumulation DCSI secondary complications in those with MODY and Novel qualifying *GCK* variants at a rate similar to that non-GCK T2D cases in both the A) UKB Full Cohort and B) Helix Cohorts. C) Hazard Ratio (HR; +/- 95%CI) to develop a DCSI score of at least 3 for each cohort, subsetted by *GCK* genotype and T2D diagnoses, compared to population controls without T2D and Hazard Ratio (HR; +/- 95%CI) for at least 2 in each DCSI secondary complication area, compared to population controls without T2D in a either cohort. Hazard ratios are generated from Cox Proportional Hazard model controlling for most recent age, sex, BMI and cohort. For display purposes, 3 categories in which there were no cases are filled in with HR of 0.1. As fewer than 5 cases were expected in these groups based on population frequencies of each phenotype and 0 cases were seen, the hazard ratios are in the range of 0.000006-0.00001 and the CIs are too large to plot well. DCSI outcomes are similar regardless of *GCK* status for the complicationI categories of peripheral vascular, cardiovascular, neuropathy and appear depleted in those with *GCK* variants for ophthalmic and cerebrovascular, and are variable by *GCK* variant group for nephropathy (too few in the metabolic category to analyze).

After confirming multi-category T2D complication trends, we next assessed if either carrier group showed enrichment or depletion for each DCSI subcategory with a reasonable case count: cardiovascular, peripheral vascular, cerebrovascular, neuropathy, neurology, and ophthalmic (retinopathy). For this analysis, we combined the clinicogenomic data from both the UKB and Helix cohorts, added an additional cohort covariate to our statistical analysis, and used age at the maximum subcategory score of 2 as the endpoint (for a UKB only analysis, see **Figure S5**). We found that MODY and Novel qualifying variant carriers with T2D diagnoses appeared just as likely as T2D cases without qualifying GCK variants to have complications classified as peripheral vascular (MODY HR=4.5, p=0.03 and Novel HR=3.1 and p=0.1 (HR=4.23 for those without *GCK* variants), cardiovascular (MODY HR=1.42, p=0.17 and Novel HR=2.0 and p=0.0001 (HR=1.75 for non-*GCK*), and neuropathy (MODY HR=2.3, p=0.25 and Novel HR=3.5, p=0.006 (HR=3.0 for non-*GCK*), and appeared less likely to develop ophthalmic or cerebrovascular complications (**Figure 4C**). Surprisingly, nephropathy-related complications had similarly significant enrichments in MODY qualifying variant carriers with T2D and T2D cases without qualifying GCK variants (MODY HR=4.3, p=0.003; non-*GCK* HR=3.4, p=0.0), while Novel qualifying variant carriers with T2D showed no enrichment against population controls without T2D (Novel HR=1.1, p=0.8).

Our retrospective analysis suggests that T2D diagnoses made in those harboring both MODY and Novel qualifying *GCK* variants are not merely misdiagnoses related to their natural elevations in blood glucose. T2D diagnosis within those harboring both *GCK* variant classes are associated with an array of T2D-related outcomes throughout the lifecourse, especially in relation to peripheral vascular, cardiovascular, and neuropathy complications. Interestingly, individuals harboring MODY qualifying *GCK* variants appear to be at particularly elevated risk not only for T2D diagnosis but also for long term nephropathy-related complications.

## Discussion

Damaging variants in *GCK* elevate blood glucose and HbA1c with high penetrance. In this work, we set out to build a population annotation strategy to globally identify all *GCK* coding variants that produce these elevations. To accomplish this we leveraged the nearly 500,000 UKB participants with exome sequencing data and random glucose measurements taken as part of a baseline health assessment. We evaluated glucose levels with regard to *GCK* variant information in a sliding window statistical analysis in training and test sets from the UKB as well as variant-level *GCK* enzymatic functional scores from a yeast-based screening assay. Next, we applied our *GCK* population genotyping strategy to the entirety of the UKB and an additional American cohort, composed of participants from three health systems (Helix cohorts). The qualifying coding *GCK* variants identified included both established pathogenic variants for MODY as well as novel variants (VUS for MODY or variants with no clinical annotation). Combined with LoFs, our *GCK* genotyping strategy identified ~1/1000 individuals from two large, unselected population cohorts carrying qualifying damaging *GCK* variants, a rate higher than previously reported using only reported pathogenic coding variants or LoFs for annotation^8^.

In line with findings from clinical studies focused on known pathogenic and LoF variants, those harboring qualifying damaging *GCK* variants using our annotation strategy have elevated glucose compared to population averages, often in the prediabetic and diabetic range, with average glucose and HbA1c levels highest in the group of individuals harboring established MODY and LoF *GCK* variants (**Figure 1C**). Aligning with the results from other gene and disease association studies, we find that T2D diagnosis risk is high for qualifying variant carriers, with novel qualifying variant carriers 3 times more likely and known pathogenic carriers 12 times more likely to be diagnosed with T2D compared to population controls and often with an earlier onset (**Figure 2**).

Somewhat unexpectedly, we also found that individuals with either established MODY or Novel damaging *GCK* variants and a T2D diagnosis have a rate of T2D-associated complications, quantified using the Diabetes Complication Severity Index (DCSI), similar to that of non-*GCK* T2D cases and nearly three times that of population controls (**Figure 4A-B**). The DCSI assesses diabetes complications in seven areas: cardiovascular, peripheral vascular, cerebrovascular, nephropathy, neurology, ophthalmic (retinopathy), and metabolic. Although the rarity of metabolic complications precluded us from completing analyses in this area, we find that certain other DCSI categories were more similar between T2D cases with and without qualifying *GCK* variants than were others (**Figure 4C**). In particular, the hazard ratio for developing cardiovascular, peripheral vascular, and neurology complications was statistically significant in T2D cases with qualifying *GCK* variants and similar to those found for T2D cases without *GCK* variants. In contrast, there appeared to be no enrichment of ophthalmic or cerebrovascular complications in T2D cases with qualifying *GCK* variants compared to the general population at all. This finding is in line with prior studies showing a lower incidence of retinopathy in those with pathogenic *GCK* variants compared to non-*GCK* T2D cases ^6,19^. Interestingly, risk for nephropathy-related complications differed by *GCK* variant group– individuals with T2D and harboring established pathogenic variants developed nephropathy complications at a rate similar to non-*GCK* T2D cases, while individuals with T2D and novel qualifying variants showed no enrichment for this phenotype. More studies are needed to confirm these trends.

On the surface, it may appear as if our results oppose other studies describing the MODY *GCK* phenotype as an isolated, stable hyperglycemia ^18,19^. Instead, it is important to consider the combination of penetrance in glucose elevations with lifetime risk of both T2D diagnosis and diabetic outcomes in individuals harboring qualifying *GCK* variants. For example, in the UKB cohort, of those harboring MODY variants, 95% have a glucose or HbA1c reading at or above prediabetes range at baseline assessment. In terms of lifetime risk, 94% have a diagnosis by age 80, but only 28% of those with MODY variants who receive a T2D diagnosis will reach a DCSI outcome of at least 3 by age 80. While this represents a substantial enrichment compared to the general population (6% to reach DCSI >3 by age 80), it means a majority of those with pathogenic qualifying *GCK* variants live their life with high glucose with few diabetic complications. This distinction is more obvious at the level of T2D diagnosis in the group of individuals with novel qualifying coding *GCK* variants, as those without a T2D diagnosis do not appear to be at any increased risk for T2D complications and instead accumulate secondary complications at the same rate as non-diabetic controls (**Figure S4**).

Taken together, those with qualifying *GCK* variants and a T2D diagnosis do develop T2D complications as measured by DCSI and would likely benefit from specific clinical care. A population screening strategy for *GCK* should consider the naturally elevated blood glucose levels across those with qualifying *GCK* variants while also appreciating that many of the patients maintain a high risk profile. For individuals with qualifying *GCK* variants and a T2D diagnosis, a more nuanced management plan may be required, as one tied to lowering glucose levels may not work for this group of patients. Follow up studies and controlled clinical trials are needed in this space.

This study was limited by the single baseline assessment with blood labs, BMI and survey questions administered in the UKB. The amount of followup also differs among participants in both the UKB and Helix cohorts. Additionally, not everyone in the Helix cohorts had glucose or HbA1c measurements available. Having measurements at multiple timepoints that started earlier in life would allow for better classification of individuals and more nuanced risk assessment. Finally, our study assessed those with known pathogenic coding and LoF GCK variants separately from other qualifying coding variants, as carriers of known pathogenic and LoF variants had significantly higher glucose and HbA1c levels. At this time, past these average glucose elevations and their known pathogenic status, the root molecular difference between coding variant carriers in the two groups is unknown, but may, at a high level, reflect different underlying molecular mechanisms of disease of damaging coding variants. For example, *GCK* MODY disease pathogenesis is typically understood via a LoF mechanism of disease, in which dampened or lowered enzymatic activity slows down overall glucose utilization and ultimately drives a higher endogenous glucose setpoint^5^. Additional molecular consequences of coding variants, such as changes that alter dynamics with interacting partners and effects on glucose uptake and glycogen regulation in the liver may also drive glucose elevations or elevated T2D risk^31,32^. A more thorough assessment of these qualifying variant groups, and follow up work in additional cohorts may help to better elucidate these patterns and ultimately could offer even more precise diagnoses and understanding of disease progression and long term complication in these carriers.

*GCK* has been lauded as a prime candidate for precision medicine. Most of the discussion has centered on using genetics to identify the subset of individuals harboring damaging *GCK* variants and stand-alone stable, mild hyperglycemia, who may not benefit or require treatment for their condition. Overall, we find that those harboring damaging *GCK* variants are at elevated risk for T2D diagnoses, and these individuals develop secondary outcomes at a rate similar to that of individuals without *GCK* variants who are diagnosed with T2D. Stand-alone glucose elevations in those harboring damaging *GCK* variants is not equivalent to a T2D diagnosis, as a subset of individuals harboring damaging variants have clinically elevated glucose without an increase in secondary complications. Population genetic screening of *GCK* may represent an important tool for both assessing future diabetes risk and ultimately making formal diabetes diagnoses, care plans targeted to specific complications of individuals with relevant variants in this gene, and providing confidence to providers to eliminate the need for anti-diabetic therapy in those for which it is not warranted.

## Methods

### Subjects and genetic data

We utilized the UK Biobank (UKB) population level exome OQFE pVCFs for 470,000 individuals (field 23157, with genotypes set to missing when DP<7 for SNVs and <10 for indels, and variants excluded if there were no homozygotes or the max allelic balance was <0.15 for SNVs or <0.2 for indels as per^33^) as well as the imputed genotypes from GWAS genotyping (field 22801-22823). The UKB study was approved by the North West Multicenter Research Ethics Committee, UK.

We also utilized 62,406 samples that were sequenced and analyzed at Helix using the Exome+^®^ assay as previously described, recruited from the Healthy Nevada Project (sequenced Jan 2018 to Mar 2023); myGenetics (sequenced May 2022 to Mar 2023); and In Our DNA SC (sequenced Dec 2021 to Mar 2023) (**Table 1**) ^34^. The Helix cohort studies were reviewed and approved by their applicable Institutional Review Boards (IRB, projects 956068-12 and 21143). All participants gave their informed, written consent prior to participation. All data used for research were deidentified.

### Phenotypes

UKB data were provided from the UKB resource (accessed September 2022). ICD codes (both ICD-9 and ICD-10) and associated dates were collected from inpatient data (category 2000), cancer register (category 100092) and the first occurrences (category 1712), which records the earliest instance of a diagnosis from the Primary Care data, Hospital inpatient data, Death Register records, and self-reported medical conditions mapped to a ICD-10 code at a three character resolution (i.e., E11 instead of E11.0). UKB glucose values (data field 30450) were converted from mmol/L to mg/dL, and HbA1c (data field 30750) were converted from mmol/mol to %. Analyses were performed both retrospectively, using all individuals with available data (Full Cohort), or prospectively, looking forward from the baseline assessment visit (between years 2006-2010) for those individuals who were both free of a T2D diagnoses as well as any self-reported use of the T2D-related drugs insulin and metformin (any of data fields 6177 “Insulin”, 6153 “Insulin”, 2986, or 20003 “insulin product or metformin”) at this time point.

Helix cohort phenotypes were processed from Epic/Clarity Electronic Health Records (EHR) data as previously described and updated as of January 2023^34^. ICD codes (both ICD-9 and ICD-10 CM) were collected from available diagnosis tables (from problem lists, medical histories, admissions data, surgical case data, account data, claims, and invoices). The data were sourced from EHR data formatted using the OMOP CDM v5.4. Each ICD source code was mapped to a source concept id and the source concept id was used to extract the relevant diagnoses.

To measure T2D-related complications, we calculated the Diabetes Complications Severity Index (DCSI), a 14-point scale calculated from ICD codes covering major complication categories^29,30^. For each of the seven DCSI categories, we used a comprehensive list of diagnosis codes including ICD9, ICD10, and ICD10CM codes (**Table S1**)^30^. For a given DCSI category, we applied the weighted score to each unique instance of the underlying diagnosis code and then transformed the total category score to 0, 1, or 2, where any summed category score over 1 was assigned a maximum score of 2. The overall DCSI score was calculated as the sum of all the transformed category scores. For the time to event analysis, we calculated age at when each DCSI score was attained (for scores 1-5), based on diagnosis records.

### Annotation and qualifying GCK variants

Variant annotation was performed with VEP-104^35^. Coding regions were defined according to Gencode version GENCODE 33, and the MANE / Ensembl canonical transcript ENST00000403799.8 was used to determine variant consequence ^36,37^. Variants from the exome were restricted to CDS (coding sequence) regions plus essential splice sites. Genotype processing for Helix data was performed in Hail 0.2.115-10932c754edb ^38^.

Variants were classified as LoF if the consequence was stop_lost, start_lost, splice_donor_variant, frameshift_variant, splice_acceptor_variant, or stop_gained and MAF<0.1% in all ancestry populations in gnomAD and the analyzed cohorts. For coding variants classified as missense_variant, inframe_deletion, or inframe_insertion, non-benign (by Polyphen or SIFT), and MAF<0.1%, we also used a combination of a functional evidence map (urn:mavedb:00000096-a), with scores for each variant assigned using a yeast complementation assay, and a statistical evidence map created using the Power Window technique to predict variants that likely increased glucose levels ^25,26,39,40^. Briefly, Power Window is a sliding window analysis that groups variants located near each other into one unit and analyzes them together to improve power, much like a gene-based collapsing analysis but at a smaller scale, as previously described ^26^. We trained the Power Window model for *GCK* on ~330k individuals (n= 333,190) from the UKB, used a beta cutoff of 0.5 to build the model, and tested on ~112k (n=112,015) unrelated UKB individuals (**Figure 1A,B**).

## Statistical analysis

Statistical analyses were run using the statsmodel package in python 3.7.3. For binary variables, logistic regression or Firth logistic regression was used; for quantitative variables, linear regression was used after rank-based inverse normal transformation; and for count data, negative binomial regression was used. For time to event analyses including cox proportional hazard calculations, the lifelines package was used, and the current age was considered to be the highest of either the age at the most recent diagnosis the participant received anywhere in their medical record, or their assessment age^41^. The main analyses were performed on all ancestries together because we are analyzing very rare variants (MAF<0.1%) collapsed together, as previously described, and the associations described perform similarly when restricted to a European-only cohort with principal components included as covariates ^13,34,42^.

## Declaration of Interests

KMSB, NT, LMM, EKB, NLW, AB and ETC are all employees of Helix. A patent has been filed by Helix for the Power Window analysis technique with ETC, KMSB, and NLW as inventors, and its current status is unpublished (application number 17575894).

## Acknowledgements

This research has been conducted using the UK Biobank Resource under Application Number 40436. Funding was provided to DRI by the Nevada Governor’s Office of Economic Development. Funding was provided to the Renown Institute for Health Innovation by Renown Health and the Renown Health Foundation. We acknowledge the entire Helix Bioinformatics team for their contributions to the production exome sequencing pipeline. We thank all of the genomic representatives of the Healthy Nevada Project, myGenetics, and In Our DNA SC (Helix Cohorts). We thank Renown Health, DRI, Medical University of South Carolina, and HealthPartners for helping to launch the Helix Cohorts.

## Data and code availability

UKB data are available for download (https://www.ukbiobank.ac.uk/) to qualified researchers. The HRN data are available to qualified researchers upon reasonable request and with permission of the HRN Steering Committee and Helix. Researchers who would like to obtain the raw genotype data related to this study will be presented with a Data Use Agreement which requires that participants will not be reidentified and no data will be shared between individuals, third parties, or uploaded onto public domains. The HRN encourages collaboration with scientific researchers on an individual basis. Examples of restrictions that will be considered in requests to data access include but are not limited to: 1. Whether the request comes from an academic institution in good standing and will collaborate with our team to protect the privacy of the participants and the security of the data requested 2. Type and amount of data requested 3. Feasibility of the research suggested 4. Amount of resource allocation for Helix and HRN member institutions required to support a collaboration.

## Funding

Funding was provided to the Desert Research Institute by the Renown Institute for Health Innovation and the Renown Health Foundation. Funding was provided to DRI by the Nevada Governor’s Office of Economic Development. Funding was provided to the myGenetics program by HealthPartners.

## Notes

### Author Declarations

The Helix cohorts were reviewed by Salus IRB (Reliance on Salus for all sites) and approved (approval number 21143), the WCG IRB (Western Institutional Review Board, WIRB-Copernicus Group) and approved (approval number 20224919), the MUSC Institutional Review Board for Human Research and approved (approval number Pro00129083), and the University of Nevada, Reno Institutional Review Board and approved (approval number 7701703417). The UK Biobank study was approved by the North West Multicenter Research Ethics Committee, United Kingdom. All participants gave their informed, written consent before participation. All data used for research were de-identified.

