## Supplemental Figures for "Patients with Type 2 Diabetes and a *GCK* variant are still at risk for T2D-related secondary complications"

**A**

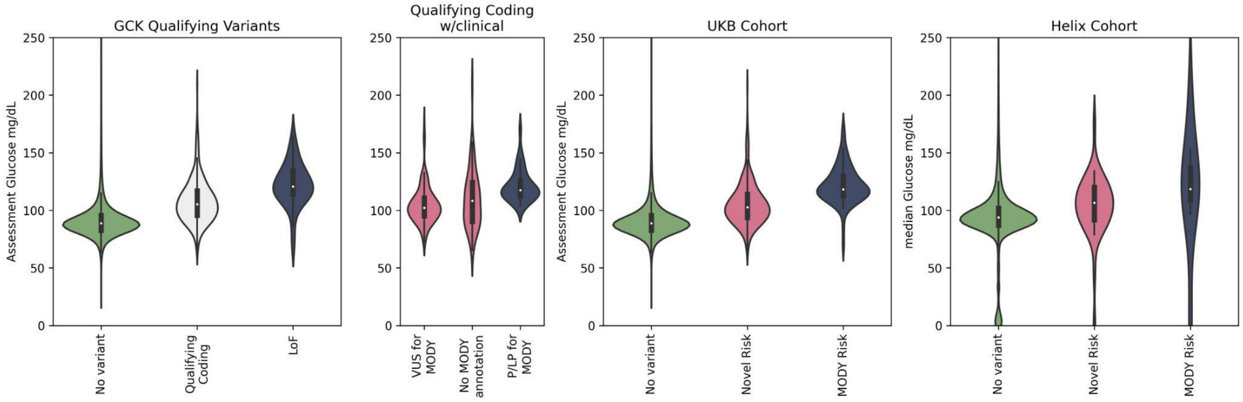


**Figure S1.** Assessment and median random glucose levels separated into the *GCK* genotypic groups used for analyses in this work, for the subset of individuals with these readings in UKB (n=397,858) and Helix (n=49,448) cohorts, respectively. Individuals with P/LP qualifying coding and LoF variants are combined for the MODY risk group and individuals harboring qualifying coding variants with either a VUS annotation or without annotation are combined for the Novel risk group (see results for details). The median random glucose levels for these groups in UKB were 89 mg/dL for non-carriers, 103 mg/dL for Novel risk group, and 118 mg/dL for MODY risk group; in Helix cohorts they were 94 mg/dL for non-carriers, 107 mg/dL for Novel risk group, and 118 mg/dL for MODY risk group. Excluded from the graph are 1641 UKB and 138 Helix participants with random glucose>=250; 53 UKB and 2 Helix participants with random glucose<40. See **Figure 1C** for HbA1c measures by genotype.

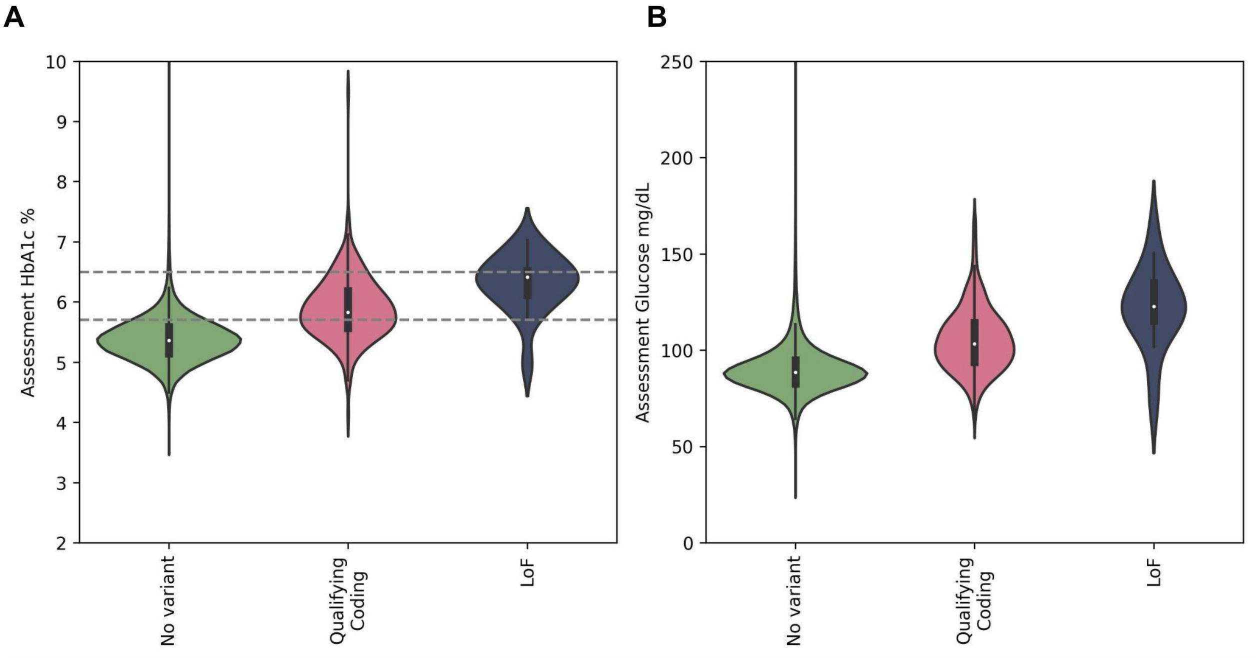


**Figure S2.** Differences in HbA1c (A) and random glucose (B) between those with qualifying coding (pink), LoF (blue), or without (green) *GCK* variants in the subset of UKB cohort who did not have a T2D diagnosis and had not reported taking common T2D medications, insulin or metformin, at the baseline assessment (Subcohort counts for HbA1c/Glucose: LoF=18/19, qualifying coding=235/238, no variant=353,764/370,675). For brevity, individuals with HbA1c values >=10 (n=269) or random glucose levels>=250 or <3 (n=374) are not shown.

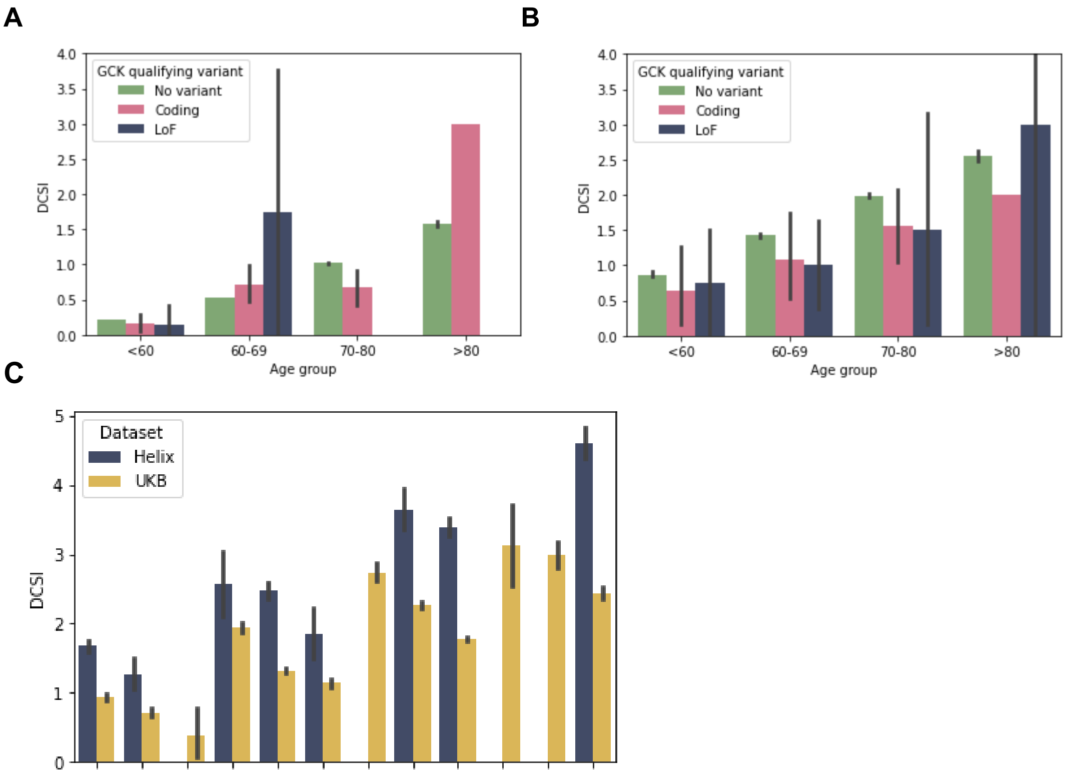


| dx age | <55 | 55-65 | >65 | <55 | 55-65 | >65 | <55 | 55-65 | >65 | <55 | 55-65 | >65 |
|---|---|---|---|---|---|---|---|---|---|---|---|---|
| Current age | <60 | <60 | <60 | 60-69 | 60-69 | 60-69 | 70-80 | 70-80 | 70-80 | >80 | >80 | >80 |

**Figure S3.** DCSI increases with age. A) UKB participants without a T2D diagnosis B) UKB participants with a T2D diagnosis. C) Helix and UKB participants split up by age at T2D diagnosis and current age. DCSI is higher at older ages and among same-aged individuals is higher if their T2D diagnosis occurred at a younger age.

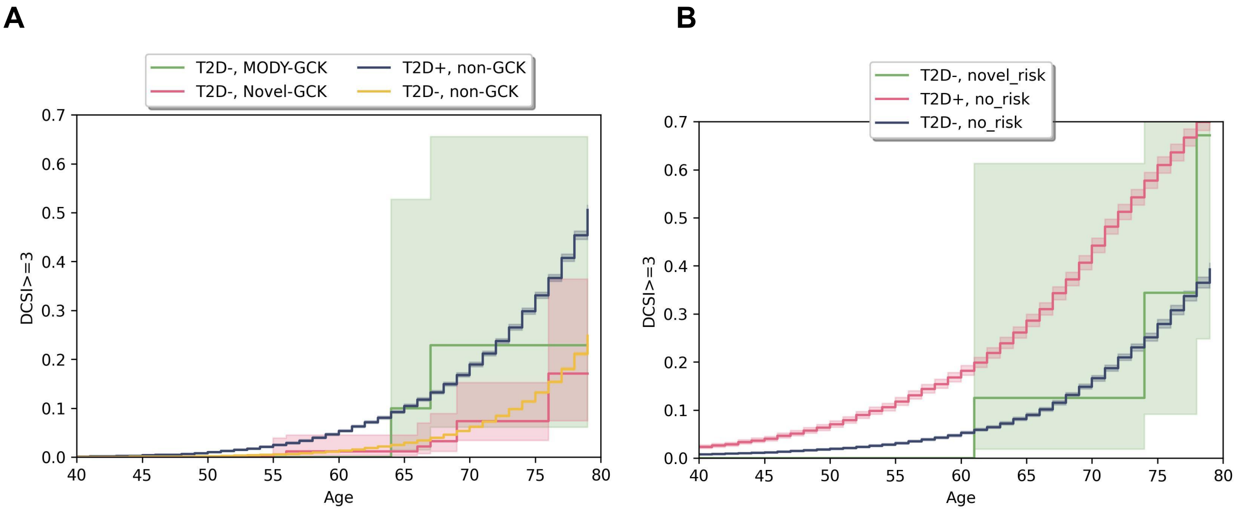


**Figure S4.** Rate of substantial DCSI complications (DCSI >=3) in MODY-*GCK* and Novel-*GCK* individuals without T2D, compared to non-*GCK* carriers with and without T2D in UKB (A) and Helix (B) cohorts. The accumulation of DCSI complications in Novel-*GCK* carriers without a T2D diagnosis are in line with that of population controls without T2D (UKB (n=228) HR= 0.81, p=0.54; Helix (n=38) HR 0.97, p=0.97). For MODY-*GCK* carriers without T2D diagnoses, statistics are not calculated due to sample size limitations (UKB n=26, Helix n=3), but time to event curve is plotted for illustrative purposes for the UKB cohort.

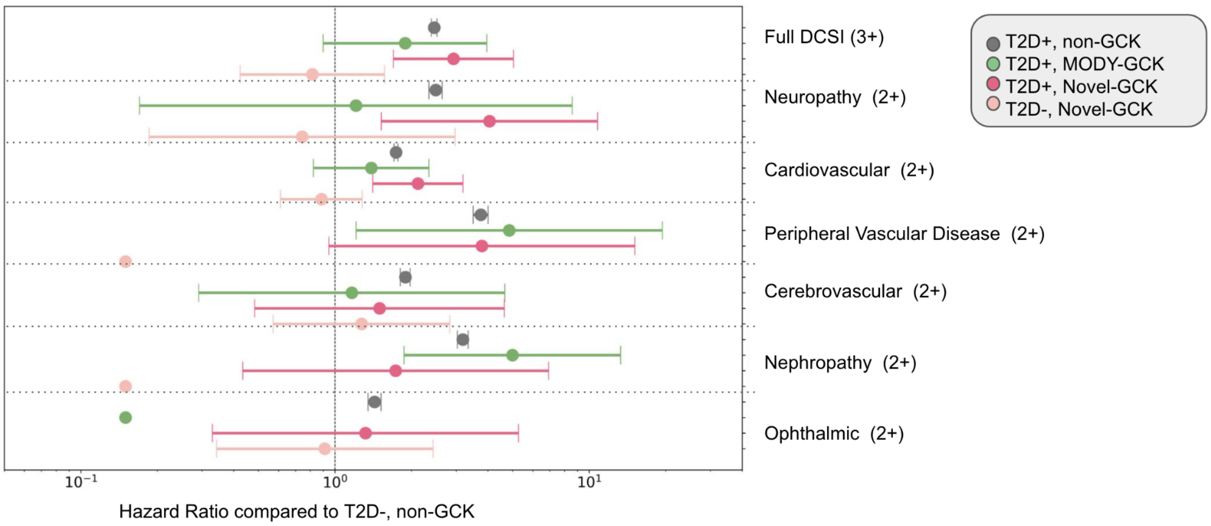


**Figure S5**. Hazard Ratio (HR; +/- 95%CI) to develop a DCSI score of at least 3 for each cohort, subsetted by *GCK* genotype and T2D diagnoses, compared to population controls without T2D and Hazard Ratio (HR; +/- 95%CI) for at least 2 in each DCSI secondary complication area, compared to population controls without T2D for the UKB Cohort. Hazard ratios are generated from Cox Proportional Hazard model controlling for most recent age, sex, BMI and cohort. For display purposes, 3 categories in which there were no cases are filled in with HR of 0.1. As fewer than 5 cases were expected in these groups based on population frequencies of each phenotype and 0 cases were seen, the hazard ratios are in the range of 0.000006-0.00001 and the CIs are too large to plot well. DCSI outcomes are similar regardless of *GCK* status for the complicationl categories of peripheral vascular, cardiovascular, neuropathy and appear depleted in those with *GCK* variants for ophthalmic and cerebrovascular, and are variable by *GCK* variant group for nephropathy (too few in the metabolic category to analyze).
